# Scanning electron microscopy observations on the surface morphology of placenta villi from HIV-positive women having preterm birth while receiving antiretroviral therapy

**DOI:** 10.1101/2023.08.11.23294011

**Authors:** Moses M. Obimbo, Shem Ochieng’, Samwel R. Gesaka, Julius Ogeng’o

## Abstract

**Background:** Preterm birth is a significant global issue. Antiretroviral therapy (ART) use and human immunodeficiency virus (HIV) infection have both been linked in recent research as independent risk factors for preterm birth. Although there has been evidence linking preterm delivery to significant pathological alterations in the placenta, it is still unclear how exactly HIV and ART harm the placenta and raise the risk of prematurity. To explain the increased risk of preterm birth (PTB), we set out to describe the surface morphological alterations in placenta villi associated with HIV and ART.

**Methods and materials:** We collected and processed 160 placentas from 40 HIV-positive women on ART and 40 HIV-negative women who had preterm deliveries, 40 HIV-positive women and 40 HIV - negative women with term delivery in Nairobi, Kenya. The placenta biopsies were harvested, washed in phosphate buffer solution, and processed for scanning electron microscopy. The dried tissue was mounted onto specimen stubs, sputter coated with gold and visualized using Zeiss Merlin FESEM in-lens. Forty representative samples, 10 from each group, were randomly selected and examined by investigators who were blinded to maternal HIV serostatus.

**Results:** The average gestational age for preterm and term births was 34 and 39 weeks, respectively. The average age of the mothers of preterm and term babies was 26.8 + 4.6 years and 24.3 + 4.3 years, respectively. The villous core of the placenta from HIV-negative patients was covered with microvilli that varied in size and appearance, and there were hardly any residual red blood cells. Placenta from HIV-positive women with preterm birth had widespread damage with shrunken and wrinkled villi, predominant blunting of the microvilli, with attendant syncytiotrophoblast disruption, and significant erythrocyte adhesion within extensive fibrillar meshwork and on surface of the syncytium.

**Conclusion:** Our results show distinctive alterations in the placenta of HIV-positive mothers who gave birth prematurely, which may impair the syncytium’s ability to function normally. Microvilli blunting, syncytial disruptions, and syncytial erythrocyte adhesion might be the symptoms of a deeper biological process. Further work to understand the effect of HIV/ART on the syncytiotrophoblast in relation to prematurity is recommended.

## Introduction

The surface interface between fetus and mother is formed by fetal syncytiotrophoblasts. These are fused multinucleated cells generated from trophectoderm and are constantly bathed in maternal blood. These cells are involved in exchange of nutrients, electrolytes, and gases between the fetus and the mother, hormone production, and are also important in fetomaternal immunotolerance [1–3]. Efficient transfer of substances across the placenta is essential in maintaining fetal growth and development and to ensure maternal health [4–6]. The surface of the placenta syncytium exposed to the maternal blood has a brush border on their free surface; This border facilitates diverse functions including absorption, secretion, and mechanotransduction [7–9]. Equally, this surface is also exposed to toxic substances and pathogens circulating in maternal blood, putting it at risk of impaired morphology [10–12].

Preterm birth (PTB) is a leading cause of perinatal morbidity and mortality in both developing and developed countries. The mechanism by which it occurs remains to be elucidated. Human immunodeficiency virus (HIV) and antiretroviral therapy (ART) have recently been identified as independent risk factors for preterm birth [13–17]. Our recent work demonstrated unfavorable histological changes in placenta from HIV positive women with preterm birth [18], similar to findings from other studies [19–21]. These findings give credence to the belief that there is a strong association between HIV and ART with adverse pregnancy outcome via diminishing placenta functional capacity. Nevertheless, the exact mechanisms by which HIV and ART cause placental damage and increase the risk of prematurity is far from being fully understood. We set out to determine the surface morphological changes on placenta villi associated with HIV and ART that could explain an increased risk of preterm labor (PTB) in this population. Scanning electron microscopy (SEM) has been used as a standard method of exploring features on the cell surface and therefore a suitable tool to use to characterize placental syncytium.

## Methods and materials

### Study setting and population

The clinical information and placental samples were collected at the Kenyatta National Hospital (KNH) and Pumwani Maternity Hospital. The two are the busiest maternity hospitals in Nairobi, Kenya each with a delivery rate of over 10,000 annually. Forty placentas were collected in the numbers given from each of the following groups: (1) HIV-positive women who received ART and had a PTB, (2) HIV-negative women who delivered preterm, (3) HIV-positive women who received ART and delivered at term, and (4) HIV-negative women who delivered at term. All the placentas were from women of African descent aged 18 to 40 years with a singleton live birth. Only women with an early pregnancy obstetric scan before 16 weeks and sure of the last normal menstrual period were included in the study. Preterm births were defined as delivery at <37 weeks of gestation and term births were defined as delivery >37 weeks of gestation. We excluded women with medical and obstetric complications during pregnancy and at the time of labor.

### Ethical considerations

Approval to conduct the study and permission to transport specimens was sought from the Kenyatta National Hospital/University of Nairobi Ethics and Research Committee (registration number P5/01/2017).

### Recruitment and informed consent

A trained research assistant cum midwife identified women who were admitted to labor and delivery meeting our inclusion criteria. He gave a full explanation of the study and obtained a written consent to enroll. Study participants were recruited from February 14, 2017, to June 22, 2017.

### Sample collection and preparation

Placentas were collected after obtaining a written consent from the recruited mothers. Six sampling sites were aseptically biopsied systematically from each placenta, trimmed and labeled a-f [22], rinsed in phosphate buffer solution and then immediately fixed in 3% glutaraldehyde solution at pH 7.2 (at ambient temperature) and stored at 4°C for 24 hours at the Kenya Aids Vaccine Initiative Institute of Clinical Research (KAVI-ICR), University of Nairobi, Kenya. The specimens were transported to the International Centre for Insect Physiology and Ecology, Kenya (ICIPE) and were post fixed in 1% osmium tetroxide in 0.1 M cacodylate buffer for 60 minutes. The blocks were then washed in distilled water three times, dehydrated in an ascending series of ethanol starting at 30%, 50%, 70%, 90% to absolute concentration for 20 minutes each and stored in epoxy/propylenoxide (1∶1) overnight. The blocks were then embedded in epoxy resin polymerized at 60°C for 36 hours and stored. Final drying was done using 100% hexamethyldisilazane (HMDS, 30mins each) and left to dry for 24hrs in a fume hood at the Central Analytical Facilities, Stellenbosch University, South Africa. Analysis of tissue ultrastructure with conventional scanning electron microscopes is labor intensive procedure and requires significant resources. To circumvent this challenge ten out of forty representative placenta samples were randomly selected from each of the above groups and examined.

### Scanning electron microscopy imaging

After mounting onto standard 12mm aluminum SEM stubs, specimens were sputter-coated with gold and visualized using Zeiss Merlin FESEM in-lens (secondary electron (SE) and SE2) detector at 1.8 mm working distance, 60 µm aperture and operated at 3-5kV keV acceleration voltage, 100 pA probe current.

## Results

A total of 160 placentas were collected and processed, 80 preterm and 80 term. All HIV-positive women were on treatment with ART. The average gestational ages of the preterm placenta was 34 (range 30-37) weeks and 39 (range 38-41) weeks for the term placenta, respectively. The mean maternal age for mothers with preterm delivery was 26.8+4.6 years and 24.3 +4.3 for the mothers with term delivery. Thirty-six (36) and 28 patients with preterm and term deliveries had cesarean deliveries, respectively.

### Structure of the placenta from HIV-negative women with preterm and term birth

The chorionic villous structures had a tree-like appearance with gradual branching. Branching seemed to increase gradually with advancing gestation. The terminal ramifications appeared as buds and some extended to the neighboring terminal villous structure via intervillous bridges (Fig 1A, B). The villous core structure was covered with microvilli of variable sizes and appearance, closely apposed against each other (Fig 1C). There were syncytial knots which appeared as dome-shaped like elevations on the surface of the syncytium and seemed to increase with advancing gestation. A few red blood cells were observed on the surface, pitting the structure of the syncytium (Fig 1D). In a few areas, the syncytial continuity was interrupted and the gap to a lesser extent filled with fibrinoid material (Fig 1E, F). There were occurrences of tip-to-tip fusion of the terminal villi in some placental sections. The preterm placenta demonstrated intermediate villi transitioning into terminal villi. In some instances, the terminal villi were cross-sectionally cut and revealed capillaries, many of which were dilated and occupied most of the cross-sectional area of the villous core (Fig 1F).

**Fig 1.**
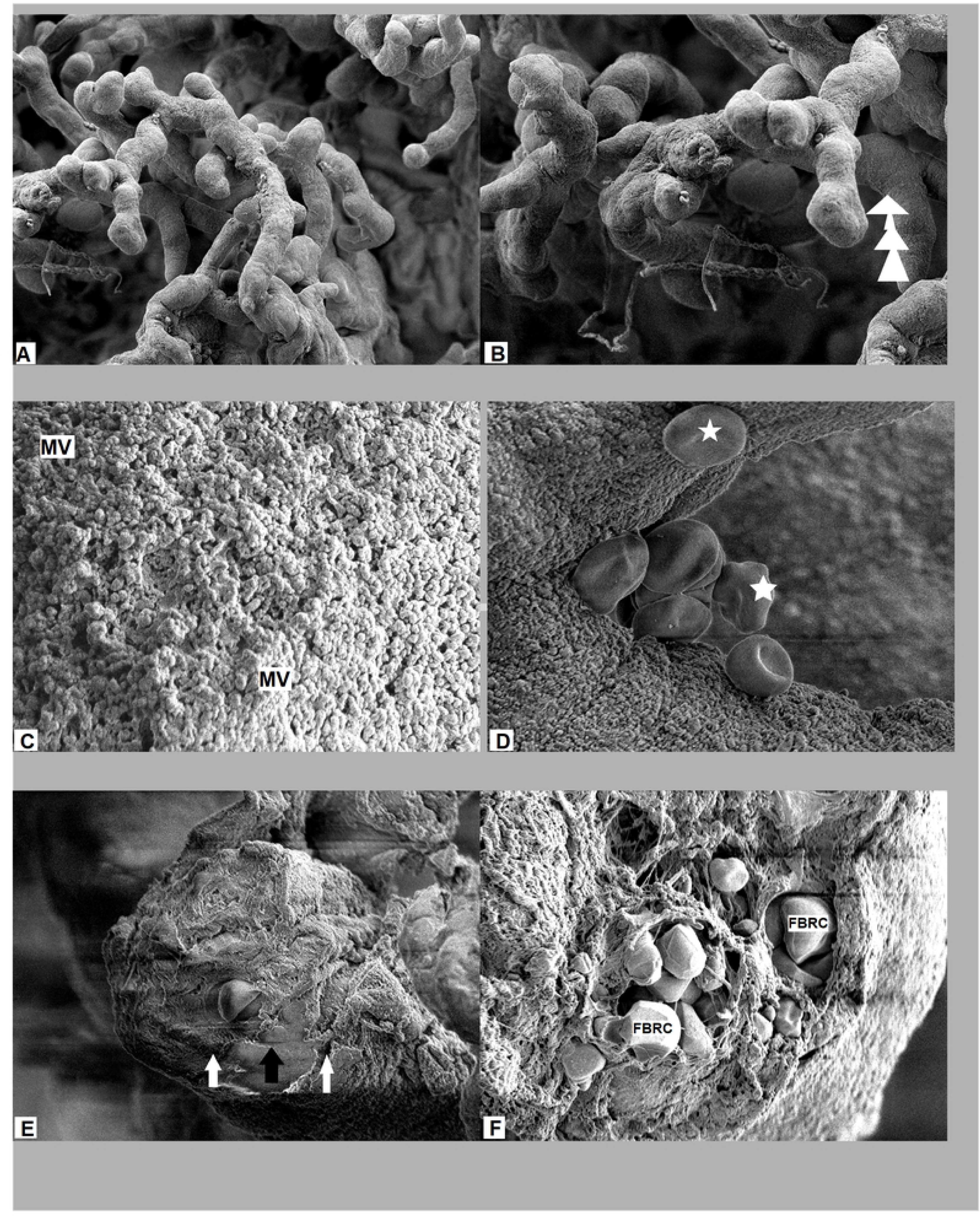
Electron microscopy structure of term and preterm placentas from HIV-negative women. (A, B) The chorionic structures with a tree-like appearance with gradual branching. The terminal ramifications and joining via intervillous bridges (white arrows). (C) The villous core structure covered with microvilli (MV) of variable sizes and appearance. (D) Red blood cells (asterisks) observed on the syncytial surface, pitting the structure of the syncytium. (E) The syncytial continuity (white arrow) disrupted and filled with fibrinoid material (black arrow). (F) Cross-sectionally cut terminal villi revealed capillaries with fetal red blood cells (FRBC).

### Structure of the placenta from HIV-positive women with preterm birth

The general structure of preterm placenta consisted of villous tree structure with intermediate and terminal villi. Most sections of these placentas showed widespread damage with shrunken and wrinkled villi. The terminal villi showed thinning with loss of congruently shaped terminal buds. Microvilli were either blunted or absent in significant areas of both the intermediate and the terminal villi with attendant syncytiotrophoblast disruption (Fig 2A, C). Massive fibrinoid material covered the cross-sectional cuts (Fig 2B). At the bases and sometimes along the surface of the terminal villi, red blood cells appeared to be adherent on the surfaces of these villi. The red blood cells were in different shapes and sizes and seemed to be trapped within extensive fibrillar meshwork (Fig 2D, E).

**Fig 2.**
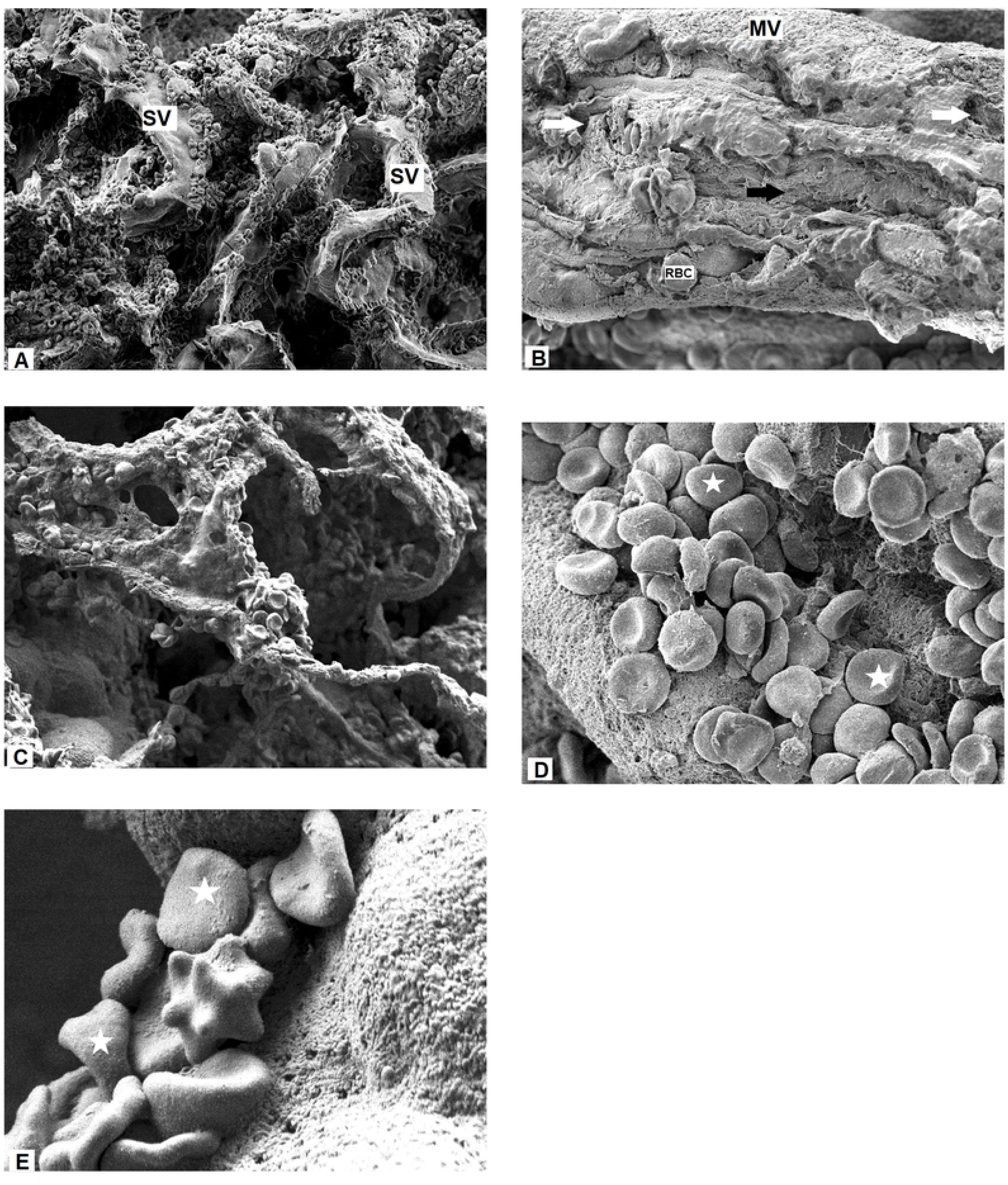
Electron microscopy structure of preterm placentas from HIV-positive women. (A) The villi appeared shrunken (SV) and wrinkled with thinning terminal villi and loss of congruently shaped terminal buds. (B) Microvilli (MV) were either blunted or absent with attendant syncytiotrophoblast disruption (white arrows). Red blood cells (RBC) were seen and massive fibrinoid material (black arrow) covered the villous structures in C, D, and E, red blood cells (asterisks) on the surface of the syncytium at advancing magnification

### Structure of the placenta from HIV-positive women with term birth

There was a progressive expansion of the villous tree with predominance of terminal villi in the term placenta. Focal villous syncytiotrophoblast thinning as a key feature was also apparent (Fig 3A). Club-shaped microvilli were present in most parts of the surface of the syncytium (Fig 3B). The red blood cells were observed on the surface of syncytium but not as significant as was seen in HIV-positive preterm placenta. The syncytium is obliterated (Fig 3C, D).

**Fig 3.**
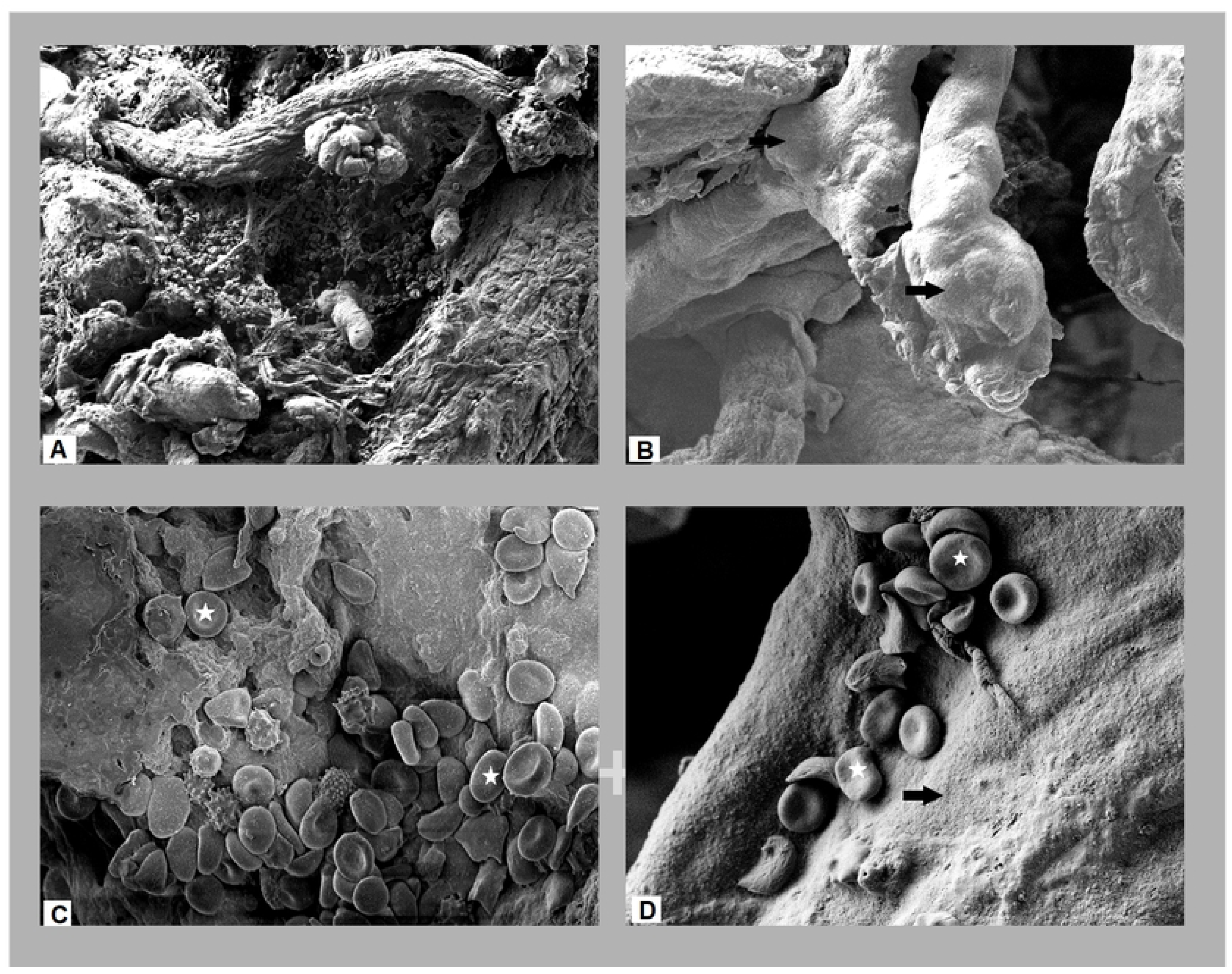
Electron microscopy structure of term placentas from HIV-positive women. (A) Focal thinning and loss of congruence of the terminal villi. (B) Club-shaped syncytial knots on the villi (black arrow). (C, D) Red blood cells trapped in the fibrillar meshwork (asterisks) on the surface of the syncytium with diminished microvilli (black arrow).

## Discussion

The maturation spectrum of the placenta from the HIV-negative women was in keeping with what has previously been observed in normal placenta. The gradual ramifications of the chorionic villi has been described and forms the functional core of the human placenta [23]. The branches are covered by the syncytialized villous trophoblast that is in direct contact with maternal blood in the intervillous space. The presence of the syncytial knots and intervillous bridges in these cases mostly signify a normal placental ageing process [24,25]. In other cases, they may be associated with pathological processes such as preeclampsia, obesity, malaria, oxidative stress and intrauterine growth restriction [26–29]. The intervillous bridges are involved in support role within placenta though the formation of an internal strut mechanism [30] and act to protect the villous capillaries from collapse during labor process. Dilatation of the capillary villi occurs at the tip of the terminal villi forming sinusoids that facilitate exchange of substances between the mother and the fetus via a thinned vasculosyncytial membranes [31].

The shrinking, wrinkling ant thinning of the placenta villi observed in placenta of HIV-positive women with preterm birth is in keeping with the decreased villi surface area and perimeter observed in our previous work [18], but has also been noted in some infections involving the placenta [32,33], intrauterine growth restriction [34], and preeclampsia [35]. By the virtue of exposure to maternal blood, the syncytium is always under constant danger of immunological or reactive oxygen species attack that could result in apoptosis, degeneration and sometimes attempt to regeneration. As stated above, the shriveling of the syncytium may be due to reduced trophoblast regeneration capacity occasioned either independently or in combination by HIV infection or the antiretrovirals. The result is that many functions of the villous syncytiotrophoblast like gaseous exchange, nutrient transfer and hormone production may be impaired.

Microvilli, found on the apical aspect of some epithelial cells are surface specialization that provide an increased surface area for absorption, secretion, mechanosensation, and also provide cellular polarity enhancing certain cellular functions such as membrane transportation, cytoskeletal distribution, and enzymatic function [8,36]. These cells, in addition, must maintain the barrier function of the compartments being separated [37]. The loss of microvilli on the surface of syncytium has been correlated with diminished placental function and has been reported in eclampsia, gestational diabetes, intrauterine growth restriction, and small for gestational age fetuses [38–40]. In the current study, the loss of microvilli may signify a possible diminished placental function which may have played a role in instigating preterm labor. Syncytiotrophoblast disruption, red cell adhesion, and an extensive fibrillar meshwork that were frequently found on the surface of the syncytium in these patients might be indicative of apoptotic process of the syncytiotrophoblast, attempt to repair, and attendant inflammatory process [41]. Fibrinoid deposition, in small amounts may be physiological as a response to minimize maternal bleeding to improve pregnancy outcome. In extensive amounts, as observed in this study, it may result in obliteration of the intervillous space, with atrophy of the chorionic villi. The basis for this observation is related to syncytiotrophoblastic damage, immunological derangement with initiation of coagulation events that results into adherence of the red blood cells on the surface of the syncytium and within the fibrinoid material [42–44]. This phenomenon heralds adverse pregnancy outcome and has been identified as a signature of maternal anti-fetal rejection [45].

The remarkable similarities illustrated in this study between the placenta from term HIV-positive patients and placenta from HIV-negative patients strongly suggest that these pregnancies were minimally affected by HIV/ART. We however, observed to a lesser extent, syncytiotrophoblast thinning, microvilli missing in some parts and the red blood cells adherence on the surface of syncytium. The difference between this group and placenta from HIV-positive women with preterm birth may be explained by interindividual biologic variability in response to drug metabolism, immune reaction or duration of HIV/ART exposure [46–48]. These results suggest HIV and ART may play a role in inducing syncytial morphological changes in placentas.

## Conclusion

Our results show distinctive alterations in the placenta of HIV-positive mothers who gave birth prematurely, which may impair the syncytium’s ability to function normally. Microvilli blunting, syncytial disruptions, and syncytial erythrocyte adhesion might be the symptoms of a deeper biological process. Further work to understand the effect of HIV/ART on the syncytiotrophoblast in relation to prematurity is recommended.

## Data Availability

All relevant data are within the manuscript and its Supporting Information files.

## Acknowledgments

All our patients who graciously donated the placenta for our research. Our research and laboratory assistants who made this work possible we say thank you.

## References

1. Ellery PM, Cindrova-Davies T, Jauniaux E, Ferguson-Smith AC, Burton GJ. Evidence for Transcriptional Activity in the Syncytiotrophoblast of the Human Placenta. Placenta. 2009.

2. Burton GJ, Jauniaux E, Charnock-Jones DS. Human Early Placental Development: Potential Roles of the Endometrial Glands. Placenta. 2007.

3. Abumaree MH, Chamley LW, Badri M, El-Muzaini MF. Trophoblast debris modulates the expression of immune proteins in macrophages: A key to maternal tolerance of the fetal allograft? Journal of Reproductive Immunology. 2012.

4. Cross JC, Werb Z, Fisher SJ. Implantation and the placenta: Key pieces of the development puzzle. Science. 1994.

5. Fisher SJ. Why is placentation abnormal in preeclampsia? Vol. 213, American Journal of Obstetrics and Gynecology. 2015. p. S115–22.

6. Salafia CM, Charles AK, Maas EM. Placenta and fetal growth restriction. Clinical Obstetrics and Gynecology. 2006.

7. Kenny AJ, Maroux S. Topology of microvillar membrance hydrolases of kidney and intestine. Physiological Reviews. 2017.

8. Lange K. Fundamental role of microvilli in the main functions of differentiated cells: Outline of an universal regulating and signaling system at the cell periphery. Journal of Cellular Physiology. 2011.

9. Al-Zuhair AGH, Ibrahim MEA, Mughal S, Abdulla MA. Loss and regeneration of the microvilli of human placental syncytiotrophoblast. Archives of Gynecology. 1987.

10. Obimbo M, Qureshi Z, Ogeng’o J. Infection in pregnancy; understanding impact on placental microenvironment and preterm birth: a review. Journal of Obstetrics and Gynecology of East and Central Africa. 2018;29(1):17–24.

11. Rudge C V., Röllin HB, Nogueira CM, Thomassen Y, Rudge MC, Odland JO. The placenta as a barrier for toxic and essential elements in paired maternal and cord blood samples of South African delivering women. Journal of Environmental Monitoring. 2009.

12. Delorme-Axford E, Sadovsky Y, Coyne CB. The Placenta as a Barrier to Viral Infections. Annual Review of Virology. 2014.

13. Ategeka J, Wasswa R, Olwoch P, Kakuru A, Natureeba P, Muehlenbachs A, et al. The prevalence of histologic acute chorioamnionitis among HIV infected pregnant women in Uganda and its association with adverse birth outcomes. Plos One. 2019;14(4):e0215058.

14. Mandelbrot L, Sibiude J. A link between antiretrovirals and perinatal outcomes? The Lancet HIV. 2017.

15. Gagnon LH, MacGillivray J, Urquia ML, Caprara D, Murphy KE, Yudin MH. Antiretroviral therapy during pregnancy and risk of preterm birth. European Journal of Obstetrics Gynecology and Reproductive Biology. 2016.

16. Cotter AM, Garcia AG, Duthely ML, Luke B, O’Sullivan MJ. Is Antiretroviral Therapy during Pregnancy Associated with an Increased Risk of Preterm Delivery, Low Birth Weight, or Stillbirth? The Journal of Infectious Diseases. 2006.

17. Nlend AEN, Motazé AN, Tetang SM, Zeudja C, Ngantcha M, Tejiokem M. Preterm birth and low birth weight after in Utero exposure to antiretrovirals initiated during pregnancy in Yaoundé, Cameroon. PLoS ONE. 2016.

18. Obimbo MM, Zhou Y, McMaster MT, Cohen CR, Qureshi Z, Ong’ech J, et al. Placental Structure in Preterm Birth Among HIV-Positive Versus HIV-Negative Women in Kenya. Journal of acquired immune deficiency syndromes (1999). 2019.

19. Castejón OC, López AJ, Pérez Ybarra LM, Castejón OC. [Placental villous lesions in HIV-1 infection treated with zidovudine]. Ginecología y obstetricia de México. 2011.

20. Shapiro RL, Souda S, Parekh N, Binda K, Kayembe M, Lockman S, et al. High prevalence of hypertension and placental insufficiency, but no in utero HIV transmission, among women on HAART with stillbirths in botswana. PLoS ONE. 2012.

21. Mohammadi H, Papp E, Cahill L, Rennie M, Banko N, Pinnaduwage L, et al. HIV antiretroviral exposure in pregnancy induces detrimental placenta vascular changes that are rescued by progesterone supplementation. Scientific Reports. 2018;8(1).

22. Burton GJ, Sebire NJ, Myatt L, Tannetta D, Wang YL, Sadovsky Y, et al. Optimising sample collection for placental research. Placenta. 2014.

23. Kingdom J, Huppertz B, Seaward G, Kaufmann P. Development of the placental villous tree and its consequences for fetal growth. European Journal of Obstetrics Gynecology and Reproductive Biology. 2000.

24. Loukeris K, Sela R, Baergen RN. Syncytial Knots as a Reflection of Placental Maturity: Reference Values for 20 to 40 Weeks’ Gestational Age. Pediatric and Developmental Pathology. 2009.

25. Fox H. THE SIGNIFICANCE OF VILLOUS SYNCYTIAL KNOTS IN THE HUMAN PLACENTA. BJOG: An International Journal of Obstetrics & Gynaecology. 1965.

26. Heazell AEP, Moll SJ, Jones CJP, Baker PN, Crocker IP. Formation of Syncytial Knots is Increased by Hyperoxia, Hypoxia and Reactive Oxygen Species. Placenta. 2007.

27. Narasimha A, Vasudeva D. Spectrum of changes in placenta in toxemia of pregnancy. Indian Journal of Pathology and Microbiology. 2011.

28. Crocker IP, Tanner OM, Myers JE, Bulmer JN, Walraven G, Baker PN. Syncytiotrophoblast degradation and the pathophysiology of the malaria-infected placenta. Placenta. 2004;

29. Roberts KA, Riley SC, Reynolds RM, Barr S, Evans M, Statham A, et al. Placental structure and inflammation in pregnancies associated with obesity. Placenta. 2011.

30. Hormann G. No Title. Archiv fur Gynakologie. 1954;184(1):109–23.

31. Jirkovská M, Janáček J, Kaláb J, Kubínová L. Three-dimensional Arrangement of the Capillary Bed and Its Relationship to Microrheology in the Terminal Villi of Normal Term Placenta. Placenta. 2008.

32. Chaikitgosiyakul S, Rijken MJ, Muehlenbachs A, Lee SJ, Chaisri U, Viriyavejakul P, et al. A morphometric and histological study of placental malaria shows significant changes to villous architecture in both Plasmodium falciparum and Plasmodium vivax infection. Malaria Journal. 2014.

33. Walter PR, Garin Y, Blot P. Placental pathologic changes in malaria: A histologic and ultrastructural study. Obstetrical and Gynecological Survey. 1983.

34. Almasry SM, Elfayomy AK. Morphometric analysis of terminal villi and gross morphological changes in the placentae of term idiopathic intrauterine growth restriction. Tissue and Cell. 2012.

35. Saleh RA, Dkhil MAM. Structural changes of placenta in preeclamptic patients: Light and electron microscopic study. Turkish Journal of Medical Sciences. 2008.

36. Lange K. Microvillar ion channels: Cytoskeletal modulation of ion fluxes. Journal of Theoretical Biology. 2000.

37. Kraehenbuhl JP, Neutra MR. Molecular and cellular basis of immune protection of mucosal surfaces. Physiological Reviews. 2017.

38. Meng Q, Shao L, Luo X, Mu Y, Xu W, Gao C, et al. Ultrastructure of Placenta of Gravidas with Gestational Diabetes Mellitus. Obstetrics and Gynecology International. 2015.

39. Salgado SS, Salgado MKR. Structural changes in pre-eclamptic and eclamptic placentas - an ultrastructural study. Journal of the College of Physicians and Surgeons Pakistan. 2011.

40. Ansari TI, Fenlon S, Pasha S, O’Neill B, Gillan JE, Green CJ, et al. Morphometric assessment of the oxygen diffusion conductance in placentae from pregnancies complicated by intra-uterine growth restriction. Placenta. 2003.

41. Roberts DJ. Placental Pathology. In: Reproductive and Developmental Toxicology. 2011.

42. Fitzgerald J, Bonnke C, Brückmann A, Schleußner E, Sossdorf M, Lösche W. Pro-coagulant capacity of syncytiotrophoblastic microparticles (STBMs). Zeitschrift für Geburtshilfe und Neonatologie. 2011.

43. Moe N, Jørgensen L. FIBRIN DEPOSITS ON THE SYNCYTIUM OF THE NORMAL HUMAN PLACENTA: EVIDENCE OF THEIR THROMBOGENIC ORIGIN. Acta Pathologica Microbiologica Scandinavica. 1968.

44. Guller S. Role of the syncytium in placenta-mediated complications of preeclampsia. Thrombosis Research. 2009.

45. Romero R, Whitten A, Korzeniewski SJ, Than NG, Chaemsaithong P, Miranda J, et al. Maternal Floor Infarction/Massive Perivillous Fibrin Deposition: A Manifestation of Maternal Antifetal Rejection? American Journal of Reproductive Immunology. 2013.

46. Yasuda SU, Zhang L, Huang SM. The role of ethnicity in variability in response to drugs: Focus on clinical pharmacology studies. Clinical Pharmacology and Therapeutics. 2008.

47. Niepel M, Spencer SL, Sorger PK. Non-genetic cell-to-cell variability and the consequences for pharmacology. Current Opinion in Chemical Biology. 2009.

48. Sweeney GD. Variability in the human drug response. Thrombosis Research. 1983.

